# Specifying uniform eligibility criteria to strengthen causal inference studies of long-term outcomes of COVID-19

**DOI:** 10.1101/2022.05.30.22275733

**Authors:** Sebhat Erqou, Andrew R Zullo, Lan Jiang, Vishal Khetpal, Julia Berkowitz, Nishant R. Shah, Justin B. Echouffo-Tcheugui, James L. Rudolph, Gaurav Choudhary, Wen-Chih Wu

## Abstract

**Background:** Causal interpretation of findings from existing epidemiological studies on long-term clinical outcomes of coronavirus disease 2019 (COVID-19) may be limited by the choice of comparator (control) group.

**Objective:** We compare two approaches to control group selection (based on requirement for negative SARS-CoV-2 test for eligibility) in long-term clinical outcomes after COVID-19 in patients with history of heart failure (HF).

**Design:** Retrospective cohort study using data from February 1, 2020 to July 31, 2021. Setting: Veteran Health Administration (VHA).

**Participants:** We studied two cohorts of Veterans with COVID-19 and history of HF which selected comparison group using two different approaches. In Cohort I, Veterans with HF who tested for positive for SARS-CoV-2 were age, sex, and race matched to Veterans with no evidence of COVID-19 in 1:5 ratio. In Cohort II Veterans with HF who tested positive for SARS-CoV-2 were age, sex, and race matched with Veterans with HF who tested negative for SARS-CoV-2 within +/-15 days of the positive test date within the same VHA facility.

**Exposure:** COVID-19 as determined by a positive SARS-CoV-2 test.

**Main Outcomes and Measures:** 1-year all-cause mortality and hospital admissions beyond the first 30 days after COVID-19 diagnosis. Adjusted hazard ratios (HRs) accounting for comorbidity and 95% confidence intervals were calculated.

**Results:** Cohort I comprised 13,722 Veterans with HF with COVID-19 (mean [SD] age 72.0 [10.2] years, 2.4% female, 71.1% White) and 60,956 matched controls not known to have COVID-19. Cohort II comprised 6,725 Veterans with HF with COVID-19 (mean [SD] age 72.5 [7.5] years, 0.1% female, 80.8% White) and 6,726 matched controls with negative SARS-CoV-2 test. The adjusted HRs for 1-year mortality and hospital admission beyond the first 30 days after diagnosis of COVID-19 were 1.40 (1.32-1.49) and 1.34 (1.28-1.41), respectively, in analysis of Cohort-I (where the comparator group was not required to test negative for SARS-CoV-2). However, in Cohort-II (using the second comparator group specifying negative SARS-CoV-2 test for eligibility), the associations were markedly attenuated; adjusted HRs 1.05 (0.95-1.17) and 1.07 (0.96-1.19), respectively.

**Conclusions:** We found significant attenuation of associations between COVID-19 and long-term risk of mortality and hospital admissions beyond the first 30 days among patient with existing HF, when comparing with a control group selected based on a negative SARS-CoV-2 test versus control group not known to have COVID-19. The findings have implications for the design of studies of long-term CVD (and non-CVD) outcome of COVID-19.

## Matter Arising (Letter Format)

We read the manuscript on long-term cardiovascular outcomes of Coronavirus disease 2019 (COVID-19) by Al-Aly et al published in Nature on 22 April, 2021, with great interest.^1^ In a retrospective cohort study using data from the Veterans Health Administration (VHA), the authors showed that “a substantial burden of health loss that spans pulmonary and several extrapulmonary organ systems is experienced by patients who survive after the acute phase of COVID-19.” The authors have also recently published another article in Nature Medicine on a wide array of long-term cardiovascular outcomes among survivors of COVID-19 using a similar approach.^2^ For these analyses, the authors sampled a comparison group of “VHA users who did not have a positive test for COVID-19”. They did not require all individuals in the comparator group to test negative for severe acute respiratory syndrome coronavirus 2 (SARS-CoV-2). Based on some of our own analyses of a smaller subset of VHA data, we believe Al-Aly et al.’s study may benefit from an additional stability analysis, specifically requiring that all individuals in the comparison group were also tested for SARS-CoV-2 and negative. This allows for the eligibility criterion of having a SARS-CoV-2 test is uniformly applied to both *exposure groups*, i.e., the *COVID-19* and the *non-COVID-19 comparator* groups.

Al-Aly et al used rigorous methods to control for confounders. They first identified a set of pre-defined and empirically identified covariates, then included these in a regression model to estimate propensity scores. These propensity scores were then used to adjust for confounding through inverse probability of treatment weights. The analytic approach to address measured confounding was sound.^3^ However, as the authors rightly acknowledge in their limitations, residual confounding remains a challenge when making causal inferences using observational data. We are concerned that the design of Al-Aly and colleagues’ study may have resulted in intractable residual (unmeasured) confounding by not requiring the *exposed (COVID-19)* and *unexposed (non-COVID-19 comparator*) groups to be exchangeable in their testing status. We suggest that those being tested for SARS-CoV-2 (whether positive or negative) are fundamentally different from the population of Veterans who are not tested. Further, we propose that only those who were tested for SARS-CoV-2 and who were negative should be sampled as the comparison group, and that time zero for all eligible Veterans should be the date of the test (or, if using discrete time, the time in which the test occurred). Al-Aly et al. implicitly required that only the COVID-19 group be tested, thus differentially applying an eligibility criterion to the exposed but not the unexposed group. The unexposed group was therefore a population of Veterans among whom an unknown fraction was tested.

Our ongoing analyses of a smaller subset of VHA data suggest that Veterans who were tested for SARS-CoV-2 were markedly different from the broader source VHA population, irrespective of whether they tested positive or negative for SARS-CoV-2. Veterans who were tested for SARS-CoV-2 may have a different distribution of socioeconomic factors (e.g., job type, income, living condition)^4,5^and behavioral factors (e.g., knowledge of prevention methods, risk behavior, health seeking behavior)^5,6^ from those who were not tested, some of which may be unmeasured. They may also have a different burden of illness that is inadequately captured by the information in the VHA databases. The data presented here provide some evidence that a better approach to address confounding may be to sample the unexposed group from among those individuals who tested negative for SARS-CoV-2, thereby applying the testing eligibility criterion uniformly regardless of exposure status. To demonstrate this, among Veterans with heart failure (HF), we identified those who developed COVID-19 between 2/1/2020 and 7/31/2021. The COVID-19 group (HF patients testing positive for SARS-CoV-2) were then compared to two different comparator groups of Veterans with HF selected using two strategies: Comparator group 1 - Veterans without a known diagnosis of COVID-19 were matched on age, sex and race to those with positive SARS-CoV-2 test, in a 1:5 ratio (‘Cohort-1’). For the exposed individuals in Cohort-1, time zero was the date of the test. For the unexposed individuals, time zero was assigned based on the distribution of the dates of time zero in the exposed group. Comparator group 2 – Veterans *who tested negative for SARS-CoV-2* were age-, sex-, race-matched to COVID-19 group in 1:1 ratio, within the same facility within +/- 15 days of the SARS-CoV-2 positive date (‘Cohort-2’). Time zero in the second comparator group was defined by the test date of SARS-CoV-2. The Comparator group 2 sampling strategy allowed us to account for geographic and time variation in the COVID-19 epidemic and, most importantly, allowed us to ensure that all individuals received a SARS-CoV-2 test regardless of their *exposure status (COVID-19 or non-COVID-19 comparator)*. Cohort 1 (based on the first comparator selection strategy) comprised 13,722 HF patients testing positive for SARS-CoV-2 and a comparator group of 60,956 HF patients with “no evidence” of COVID-19. Cohort-2 (based on the second comparator selection strategy) comprised of 6725 HF patients testing positive for SARS-CoV-2 and a comparator group of 6766 HF patients who definitively tested negative for SARS-CoV-2 (**Table**).

**Table.**
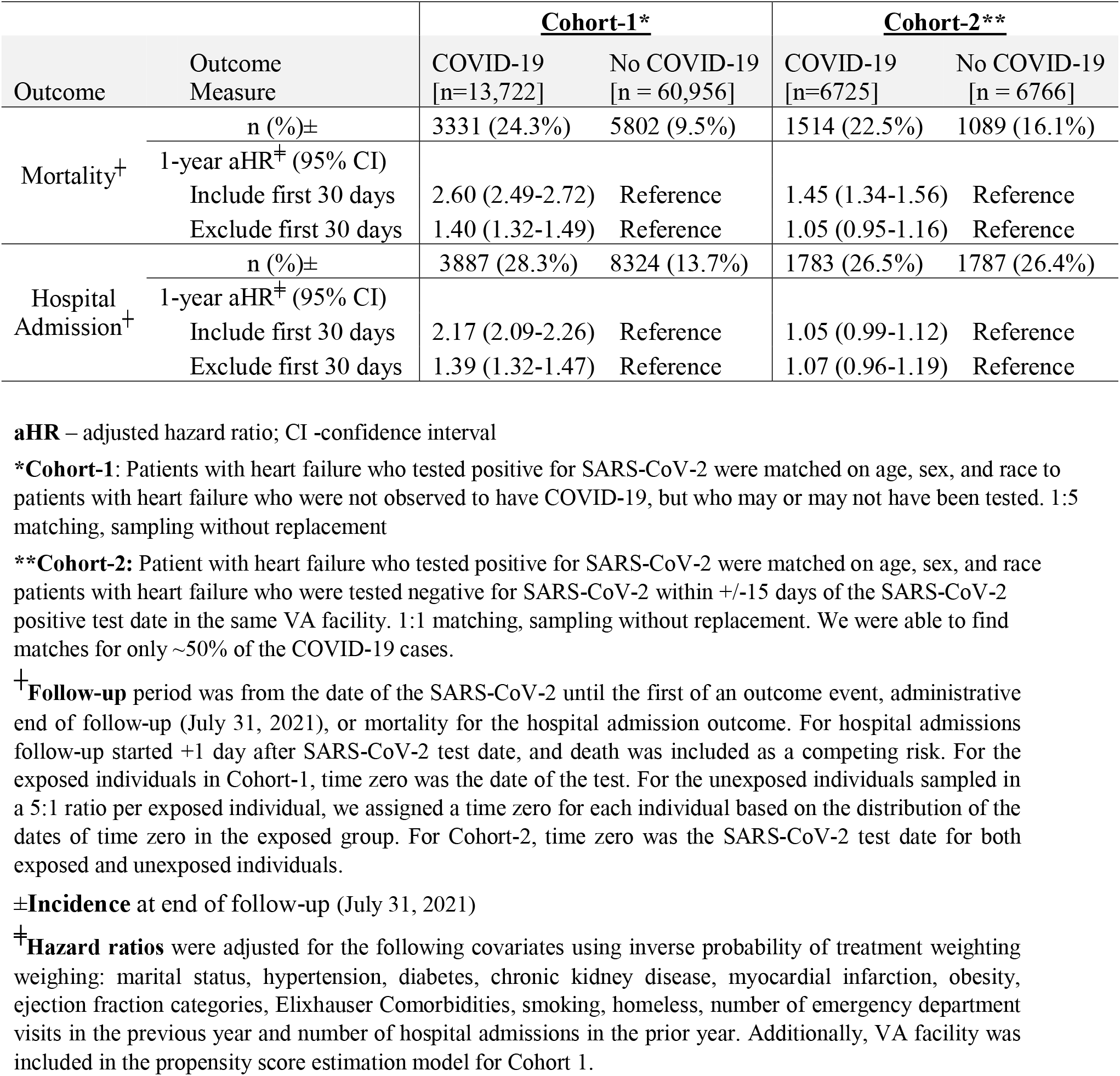
Long-term incidences and hazard ratios for COVID-19 mortality and hospital admission outcomes among Veterans with heart failure when using two approaches to sampling the comparator (non-COVID-19) group

As shown in the **Table**, the adjusted hazard ratios (HRs) for 1-year mortality and hospital admission beyond the first 30 days after diagnosis of COVID-19 were 1.40 (1.32-1.49) and 1.34 (1.28-1.41), respectively, when using the first comparator group (i.e., analysis of Cohort-1). However, in Cohort-2, the associations were markedly attenuated – adjusted HRs 1.05 (0.95-1.17) and 1.07 (0.96-1.19), respectively. This may suggest the presence of residual confounding in the first set of analyses despite adjusting for a wide array of covariates, possibly due to unmeasured or inaccurately measured confounders. As shown in the **Figure** the standardized mean differences (SMDs) comparing covariates between the COVID-19 group and the comparator group were all ≤0.1 for Cohort-2 except for EF categories (SMD = 0.11) while for Cohort-1, 14 variables had SMD >0.1, indicating that the exposed and unexposed groups were more balanced in terms of the measured covariates in Cohort-2. As these covariates are known to be associated with the outcome of interest, this suggests that in Cohort-2 confounding may have been mitigated through the study design decision to require all individuals (both exposed and unexposed) to have received a SARS-CoV-2 test. Those tested for SARS-CoV-2, even when testing negative, had higher comorbidity burden (***Supplement Table***), and had higher hospitalization and mortality rates (**Table**) compared to those who were not tested. It is possible that differences in unmeasured or inadequately measured covariates may follow the same pattern as observed for measured covariates.

**Figure.**
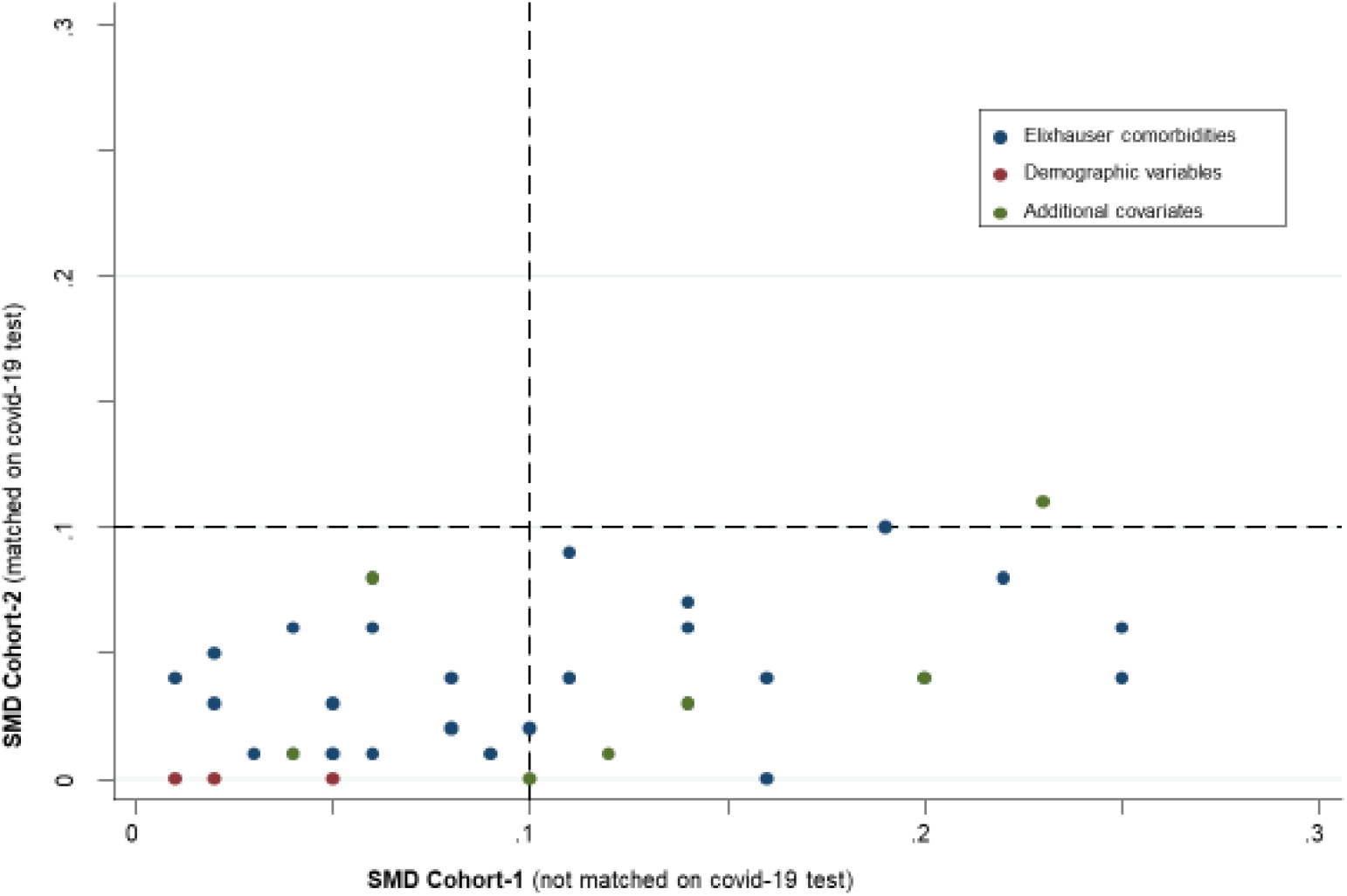
Plot of standardized mean differences in covariates comparing the exposed (COVID-19) and unexposed (non-COVID comparator) groups in the two cohorts using two different approaches to sampling the non-COVID comparator group for Veterans with heart failure. **SMD** – standardized mean difference **\*Cohort-1**: Patients with heart failure who tested positive for SARS-CoV-2 were matched on age, sex, and race to patients with heart failure who were not observed to have COVID-19, but who may or may not have been tested. 1:5 matching, sampling without replacement. **\*\*Cohort-2:** Patient with heart failure who tested positive for SARS-CoV-2 were matched on age, sex, and race patients with heart failure who were tested negative for SARS-CoV-2 within +/-15 days of the SARS-CoV-2 positive test date in the same VA facility. 1:1 matching, sampling without replacement. We were able to find matches for only ∼50% of the COVID-19 cases. **Elixhauser comorbidities** - include 29 variables **Demographic variables** - age, sex, race (both cohorts matched for these variables) **Additional covariates** - smoking, homelessness, myocardial infarction, chronic kidney disease, chronic obstructive lung disease, heart failure, Ejection Fraction (EF) categories (<45%, ≥45%, missing) **Note:** all SMDs were ≤0.1 for Cohort-2 except for EF categories (SMD = 0.11) compared to Cohort-1 where 14 variables had SMD >0.1, indicating that the exposed and unexposed individuals were more balanced in terms of the measured covariates in Cohort-2.

Sampling exposed and unexposed individuals with the same eligibility criteria, including SARS-CoV-2 testing status, appears to potentially account (through design) for some residual confounding. In addition, specifying negative SARS-CoV-2 testing for eligibility to comparator group is important to reduce bias arising from misclassification of the exposure. It is estimated that at least 40% SARS-CoV-2 infections are asymptomatic,^7^ and since the comparator group inclusion criteria by Al-Aly et al did not require a negative for SARS-CoV-2, these asymptomatic SARS-CoV-2 cases would be included in their study as controls resulting in non-random misclassification of the exposure. If one makes a reasonable assumption that individuals with asymptomatic SARS-CoV-2 infection are less likely to develop long-term sequala (compared to those with symptomatic infection), then this non-random misclassification will cause a bias away from the null resulting in associations that are stronger than the “true” relationship existing in the population. The assumption that asymptomatic SARS-CoV-2 infections (compared to symptomatic infections) are less likely to be associated with long-term clinical outcomes is consistent with the observation by Al-Aly et al of a graded increase in hazard ratios of long-term outcomes comparing COVID-19 cases who, (i) were not hospitalized, (ii) were hospitalized, or (iii) were admitted to intensive care unit, with their control group, as shown in Figure 3 of their article in Nature.^1^

Another explanation for the differences in the results between the two approaches might be differences in the distributions of effect measure modifiers introduced by selecting a different comparator group. Hence, the differences in the estimates that we observed may not necessarily represent an improved confounding control; rather, it may represent different overall distributions of effect measure modifiers between the cohorts. It is also possible that our sampling strategy of selecting the comparator individuals within the same facility and within the same fine strata of calendar time as the individuals with COVID-19, rather than our requirement that both the exposed and unexposed group should meet the same eligibility SARS-CoV-2 testing criterion, could be more responsible for any reductions in residual confounding or selection bias.

Nonetheless, when studying long-term COVID-19 outcomes, we believe the approach demonstrated here of sampling the unexposed comparator individuals from among those tested for SARS-CoV-2 at the same time and place as the COVID-19 exposed group allows the exposed and unexposed groups to be more exchangeable in their covariate distributions even before any statistical adjustment. We hope that Al-Aly et al. will consider this additional stability analysis to shed additional light on their findings. A similar comparator group selection strategy has been implemented in the design of at least one other currently on-going study of long-term CVD outcomes among survivors of COVID-19.^8^ If the results of Al-Aly et al. persist after applying the eligibility criteria (including SARS-CoV-2 testing) to both exposed and unexposed groups uniformly, that information will be a very important contribution to scientific knowledge. If the results are not robust to this change, that will guide future investigators to avoid sampling a mixture of individuals who were and were not tested for SARS-CoV-2 as an unexposed comparator group.

## Data Availability

Based on restrictions in the Data Use Agreements used in this study, the authors are unable to make a data set available. Methodology questions may be directed to the corresponding author.

## Acknowledgement

The reported/outlined here was supported by the Department of Veterans Affairs, Veterans Health Administration, VISN 1 Career Development Award to SE. SE is also funded by Center for Aids Research, The Rhode Island Foundation, and Lifespan Cardiovascular Institute. JLR, WW, SE, and LJ were supported by the VA Health Services Research and Development Center of Innovation in Long Term Services and Supports (CIN 13-4193; C19-20-213). ARZ was supported in part by grants from the National Institute on Aging (R21AG061632, R01AG065722, RF1AG061221, R24AG064025, and R01AG062492). GC was supported by NIGMS P20GM103652, VA CSR&D grant I01CX001892, and NHLBI R01HL148727. The views expressed in this article are those of the authors and do not necessarily reflect the position or policy of the Department of Veterans Affairs or the United States government.

---

**Supplement Table.** Comorbidity burden in the two comparator groups for Veterans with heart failure identified using two selection strategies (comparator group-1 selected among individuals not known to have COVID-19, comparator group-2 selected among individuals who tested negative for SARS-CoV-2).

| **Variable** | **Comparator Group-1**<br>**Mean (SD) or n (%)** | **Comparator Group-2:**<br>**Mean (SD) or n (%)** | **SMD** |
| --- | --- | --- | --- |
| Age (years) | 72.0 (10.2) | 72.5 (7.5) | -0.06 |
| Elixhauser comorbidity index | 6.1 (3.0) | 7.5 (3.1) | -0.44 |
| Female | 1473 (2.4%) | * (0.1%) | 0.20 |
| Race-White | 43336 (71.1%) | 5469 (80.8%) | 0.34 |
| Race-Black | 13281 (21.8%) | 1191 (17.6%) |  |
| Chronic kidney disease | 18709 (30.7%) | 2570 (38.0%) | -0.15 |
| Homeless | 1899 (3.1%) | 236 (3.5%) | -0.02 |
| Myocardial infarction | 33481 (54.9%) | 4351 (64.3%) | -0.19 |
| Smoker | 11283 (18.5%) | 1740 (25.7%) | -0.17 |
| Chronic obstructive lung disease | 21564 (35.4%) | 3046 (45.0%) | -0.20 |
| Valvular disease | 9014 (14.8%) | 1365 (20.2%) | -0.14 |
| Pulmonary circulation disorders | 4687 (7.7%) | 765 (11.3%) | -0.12 |
| Peripheral vascular disorders | 17000 (27.9%) | 2633 (38.9%) | -0.24 |
| Paralysis | 622 (1.0%) | 104 (1.5%) | -0.05 |
| Neurodegenerative disorders | 5771 (9.5%) | 909 (13.4%) | -0.12 |
| Chronic pulmonary disease | 21773 (35.7%) | 3085 (45.6%) | -0.20 |
| Diabetes without complications | 25936 (42.5%) | 3253 (48.1%) | -0.11 |
| Diabetes with complications | 24089 (39.5%) | 3304 (48.8%) | -0.19 |
| Hypertension without complications | 47531 (78.0%) | 5720 (84.5%) | -0.17 |
| Hypertension with complications | 23689 (38.9%) | 3298 (48.7%) | -0.20 |
| Cardiac arrhythmias | 33304 (54.6%) | 4287 (63.4%) | -0.18 |
| Hypothyroidism | 7640 (12.5%) | 960 (14.2%) | -0.05 |
| Renal Failure | 18856 (30.9%) | 2581 (38.1%) | -0.15 |
| Liver disease | 4770 (7.8%) | 747 (11.0%) | -0.11 |
| Peptic ulcer disease, no bleeding | 514 (0.8%) | 90 (1.3%) | -0.05 |
| AIDS/ HIV | 304 (0.5%) | 55 (0.8%) | -0.04 |
| Lymphoma | 855 (1.4%) | 134 (2.0%) | -0.04 |
| Metastatic cancer | 776 (1.3%) | 186 (2.7%) | -0.11 |
| Solid tumor without metastasis | 6475 (10.6%) | 1096 (16.2%) | -0.16 |
| Rheumatoid arthritis/collagen vascular diseases | 1745 (2.9%) | 293 (4.3%) | -0.08 |
| Coagulopathy | 2381 (3.9%) | 421 (6.2%) | -0.11 |
| Obesity | 15563 (25.5%) | 2031 (30.0%) | -0.10 |
| Weight loss | 2904 (4.8%) | 490 (7.2%) | -0.10 |
| Fluid and electrolyte disorders | 11267 (18.5%) | 1951 (28.8%) | -0.25 |
| Blood loss anemia | 6207 (10.2%) | 1064 (15.7%) | -0.17 |
| Deficiency anemia | 966 (1.6%) | 190 (2.8%) | -0.08 |
| Alcohol use | 5322 (8.7%) | 744 (11.0%) | -0.08 |
| Drug use | 3464 (5.7%) | 421 (6.2%) | -0.02 |
| Psychosis | 1438 (2.4%) | 188 (2.8%) | -0.03 |
| Depressions | 15393 (25.3%) | 2108 (31.2%) | -0.13 |
| Ejection Fraction $\geq 45\%$ | 23703 (38.9%) | 2834 (41.9%) | 0.39 |
| Ejection Fraction $< 45\%$ | 24968 (41.0%) | 3440 (50.8%) | |

SMD – standardized mean difference.
**Comparator group-1:** Patients with heart failure who tested positive for SARS-CoV-2 were matched on age, sex, and race to patients with heart failure who were not observed to have COVID-19, but who may or may not have been tested. 1:5 matching, sampling without replacement

## References

1. Al-Aly, Z., Xie, Y. & Bowe, B. High-dimensional characterization of post-acute sequelae of COVID-19. Nature 594, 259–264 (2021). https://doi.org/10.1038/s41586-021-03553-9

2. Xie, Y., Xu, E., Bowe, B. et al. Long-term cardiovascular outcomes of COVID-19. Nat Med (2022). https://doi.org/10.1038/s41591-022-01689-3

3. Lunceford JK and Davidian M. Stratification and weighting via the propensity score in estimation of causal treatment effects: a comparative study. Stat Med (2004). 23(19):2937–60. https://doi.org/10.1002/sim.1903

4. Ahmad K, Erqou S, Shah N, Nazir U, Morrison AR, Choudhary G, et al. Association of poor housingconditionswithCOVID-19incidenceand mortality across US counties. PLoSONE 15(11):e0241327 (2020). https://doi.org/10.1371/journal.pone.0241327

5. Khanijahani A, Iezadi S, Gholipour K, et al. A systematic review of racial/ethnic and socioeconomic disparities in COVID-19. Int J Equity Health. 20(1):248 (2021). https://doi:10.1186/s12939-021-01582-4

6. Alsan M, Stefanie Stantcheva S, Yang D, et al. Disparities in Coronavirus 2019 Reported Incidence, Knowledge, and Behavior Among US Adults. JAMA Netw Open. 3(6):e2012403 (2020). https://doi:10.1001/jamanetworkopen.2020.12403

7. Oran DP, Topol EJ. Prevalence of Asymptomatic SARS-CoV-2 Infection : A Narrative Review. Ann Intern Med. 2020 Sep 1;173(5):362–367. doi: https://10.7326/M20-3012

8. Arévalos V, Ortega-Paz L, Fernandez-Rodríguez D, et al. Long-term effects of coronavirus disease 2019 on the cardiovascular system, CV COVID registry: A structured summary of a study protocol. PLoS ONE 16(7): e0255263 (2021). https://doi.org/10.1371/journal.pone.0255263

